# The Transdiagnostic Association between Cognitive Functioning and Psychopathology *Exploratory Modeling of Cognitive Structure in a Naturalistic Patient Sample*

**DOI:** 10.64898/2026.02.03.26345448

**Authors:** J.D. Kist, J.N. Vrijsen, C. Fraza, R.M. Collard, P.C.R. Mulders, A. Marquand, I. Tendolkar, P.F.P. van Eijndhoven

## Abstract

**Background:** Impairments in cognitive functioning (CF) contribute to the onset, severity, and persistence of psychiatric symptoms. While specific CF domains may relate differentially to psychopathology, evidence also supports a general factor of cognitive impairment (the C-factor). We aimed to examine how general and domain-specific CF impairments relate to psychopathology using both diagnosis-specific and transdiagnostic symptom frameworks.

**Methods:** Data were drawn from five cognitive tasks administered in the deep-phenotyped, naturalistic MIND-Set cohort. A bifactor model of CF was estimated in a discovery sample (n = 206) and internally validated in a separate subsample (n = 312). Factor scores were then explored in relation to broad diagnostic clusters (stress-related disorders, neurodevelopmental disorders, comorbid disorders, and healthy controls), presence of specific diagnoses, number of diagnoses, and transdiagnostic symptom domains.

**Results:** The bifactor model comprised a general CF factor (C-factor) and five specific subfactors—Reaction Time, Incompatibility, Working Memory, Inhibition, and Flexibility—and successfully replicated, although the general factor was relatively weak. Diagnosis-specific analyses showed that only individuals with stress-related disorders differed significantly from healthy controls on the C-factor and the Incompatibility factor. Higher impairment on the Incompatibility factor was associated with mood disorder diagnoses, while both the C-factor and Incompatibility factor were correlated with greater diagnostic burden. At the symptom level, the Incompatibility factor was associated with Negative Valence and Arousal domains, the C-factor with Negative Valence, and the Flexibility factor with Arousal.

**Conclusion:** These findings indicate that broader cognitive impairment and deficits on tasks requiring inhibition under cognitive load are primarily related to mood disorders, ADHD, and transdiagnostic symptoms of negative valence and arousal. More generally, cognitive impairment appears to reflect symptom burden and transdiagnostic expression rather than diagnostic category alone, suggesting that dimensional symptom measures may provide a more informative framework for understanding cognitive impairment in clinical populations.

## Background

Psychiatric patients often exhibit broad impairments in cognitive functioning (CF), which can contribute to the onset, persistence, and severity of symptoms. Deficits span attention, memory, processing speed, and higher-order control processes, each of which undermines daily, social, and occupational functioning (Letkiewicz et al., 2014; Snyder, Miyake, & Hankin, 2015). Understanding the nature and structure of these impairments is crucial for understanding the relationship with daily functioning, symptom severity and mental health in general. While cognitive deficits are common across many forms of psychopathology, the type and extent of impairment may vary depending on the disorder (Snyder et al., 2015). For example, impairments in inhibitory control have been mostly linked to Attention Deficit and Hyperactivity Disorder (ADHD), while for Autism Spectrum Disorder (ASD) and anxiety this is mainly to impairments in flexibility (Craig et al., 2016; D’Cruz et al., 2013; Lee & Orsillo, 2014).

Executive functions (EF) are higher-order processes—cognitive inhibition, mental flexibility, and working memory—that regulate and organize more basic cognitive operations (East-Richard, R.-Mercier, Nadeau, & Cellard, 2020; Waserstein et al., 2019). While EF is central to adaptive behavior, it represents only one dimension of CF and may not capture all deficits observed across disorders. Studying CF domains in isolation proves difficult due to their interdependence and the high rates of comorbidity and heterogeneity in psychiatric samples (Bloemen et al., 2018; Dajani, Llabre, Nebel, Mostofsky, & Uddin, 2016; van Hal et al., 2023). Focusing on single domains and single diagnoses risks overlooking shared variance and transdiagnostic deficits that cut across traditional diagnostic boundaries.

Dimensional models introduce a general cognitive factor (the C-factor) that reflects shared impairments across disorders (Abramovitch, Short, & Schweiger, 2021; Bloemen et al., 2018; East-Richard et al., 2020). Similar to the P-factor and G-factor, which describe a general level of psychopathology and general intelligence respectively, the C-factor is a converging factor that simplifies the complex, multidimensional concept of cognitive functioning (Caspi et al., 2014; Pettersson, Larsson, D’Onofrio, Bölte, & Lichtenstein, 2020). A bifactor model provides an elegant solution for mapping CF impairments, offering a framework for understanding how this global deficit factor interacts with domain-specific weaknesses (Gegenfurtner, 2022). It estimates a general factor alongside orthogonal domain-specific factors (e.g., flexibility, working memory), clarifying how much variance each source explains within complex, intercorrelated data.

This paper reports findings from a naturalistic patient sample that includes individuals with stress-related disorders, neurodevelopmental conditions, and combinations of these disorders, as well as non-disordered controls. All participants completed a battery of computerized tasks designed to measure multiple CF domains. We sought to determine whether a bifactor model could elucidate the latent structure of CF impairments and relate those factors to both traditional diagnoses and transdiagnostic symptom dimensions.

We therefore formulated the following three research aims: **(I)** To develop and evaluate a bifactor model of cognitive functioning across multiple domains, capturing both a general factor and domain‐specific factors using neurocognitive data from a naturalistic patient population. **(II)** To examine the relationship between general and domain-specific factors of CF and traditional diagnoses. This includes identifying how different groups of psychiatric diagnoses differ between, and are associated with, factors of CF. Additionally, we will test whether the impairment in these factors of CF is associated with the number of diagnoses an individual has. **(III)** To explore associations between CF and transdiagnostic measures of psychopathology. This involves assessing how the factors of CF relate to symptom dimensions that cut across traditional diagnostic boundaries.

## Methods

### Participants and Diagnoses

This study includes 518 participants from the MIND-Set (*Measuring Integrated and Novel Dimensions*) cohort. The participant sample consists of patients (N = 495) who were referred to the outpatient clinic of the Psychiatry department of the Radboudumc in Nijmegen, the Netherlands, and non-disordered controls (N = 123). Patients could receive a diagnoses of a mood-related disorder, addiction, anxiety-related disorder, autism spectrum disorder (ASD) and attention-deficit hyperactivity disorder (ADHD). Diagnoses were confirmed by trained clinicians using the Structured Clinical Interview for DSM-IV Axis I Disorders (SCID-I) (REF). Inclusion of the control sample was dependent on the absence of a current or past psychiatric disorder (van Eijndhoven et al., 2022).

Patients who were diagnosed with a stress-related disorder, such as depression or anxiety, can be grouped into the stress-related (SR) group. Patients who received a diagnoses of ASD or ADD or both, into the neurodevelopmental (ND) group. Patients who were diagnosed with both a stress-related and neurodevelopmental disorder were categorized in the comorbidity (CD) group. Patients who were diagnosed solely with addiction were not included in this sample, while addiction was included with patients as a comorbid disorder. The number of diagnoses one individual could have therefore ranges from zero to five, potentially including a mood-disorder, anxiety disorder, ADHD, ASC and addiction.

### Measures

#### Dimensional Symptom Severity

Symptom severity was measured using validated self-report questionnaires. These included questionnaires specific to disorders, such as depression (Inventory of Depressive Symptomatology, IDS-SR), anxiety (Anxiety Sensitivity Scale, ASI), ASD (Autism-Spectrum Quotient (Dutch version), AQ-50NL) and ADHD (Connor’s Adult ADHD Rating Scale, CAARS) (Baron-Cohen, Wheelwright, Skinner, Martin, & Clubley, 2001; Conners, 1999; Rush, Gullion, Basco, Jarrett, & Trivedi, 1996; Taylor et al., 2007). Furthermore, questionnaires measuring subjective cognitive performance (Brief Assessment of Impaired Cognition Questionnaire (BRIEF-A)), personality (Personality Inventory for DSM-IV Short Form (PID)), perseverative thinking (Perseverative Thinking Questionnaire (PTQ) and alexithymia (Toronto Alexithymia Scale (TAS-20)) were set out for self-report (Al-Dajani, Gralnick, & Bagby, 2016; Bagby, Parker, & Taylor, 1994; E. Scholte, 2011; Ehring et al., 2011).

The transdiagnostic factor structure applied in the present study was originally conceptualized in Mulders and colleagues (2022). This exploratory factor analyses uses the above symptom-measures. The resulting factors closely resembled four domains of the RDoC, namely *Negative Valence, Social Systems, Cognition* and *Arousal/Inhibition* (Insel, 2014). The same exploratory factor analysis was conducted on the present participant sample as described in detail in Kist et al. (2025), which reports the full analytic procedure and results. The factor structure explains 68.4% of the variance of the data and has a Kaiser-Meyer-Olkin measure of 0.956 and a significant Bartlett’s test of Sphericity (p<0.001).

#### Cognitive functioning

Participants underwent a neurocognitive assessment, which included three tasks from the Test of Attentional Performance (TAP) (version 2.3), two tasks from the Cambridge Neuropsychological Test Battery (CANTAB) and the Probabilistic Reversal Learning (PRL) Task (see supplementary materials table 1) (Cambridge Cognition; Cools, Barker, Sahakian, & Robbins, 2001; Swainson et al., 2000; Zimmermann & Fimm, 2002). Within the TAP battery, the main aspects of CF that are measured include basal responsiveness, cognitive flexibility, and inhibition. The different tasks of the CANTAB assess spatial working memory and cognitive flexibility. Finally, the PRL tests probabilistic reasoning, reversal learning, and reward processing.

**Table 1.** Descriptives of MIND-Set Participant Samples. Participant dataset was split into a Discovery sample: based on the availability of only data on Cognitive Functioning (CF) and a Replication Sample, including participants with both CF and Magnetic Resonance Imaging (MRI) data. Disorder Categories includes the categories stress-related, patients with a diagnoses of depression and/or anxiety, neurodevelopmental: patients with a diagnosis of autism spectrum disorder and/or attention deficit and hyperactivity disorders, and comorbid, patients with at least one stress related and one neurodevelopmental disorders.

|  | Discovery<br>Sample | Replication<br>Sample | Total |
| --- | --- | --- | --- |
| **N** | 206 | 312 | 518 |
| **Disorder Categories** |  |  |  |
| Stress-related<br>disorder<sup>1</sup> | 77 (37.4%) | 90 (28.8%) | 167 (32.2%) |
| Neurodevelopmental<br>disorder | 16 (7.8%) | 23 (7.4%) | 39 (7.5%) |
| Comorbid Disorders | 75 (36.4%) | 114 (36.5%) | 189 (36.5%) |
| Control | 38 (18.4%) | 85 (27.2%) | 123 (23.7%) |
| **Age** |  |  |  |
| Mean (SD) | 42.99<br>(15.11) | 37.646<br>(14.52) | 39.774 (14.97) |
| Range | 18.62 -<br>79.48 | 17.92 -<br>74.98 | 17.92 - 79.48 |
| **Sex** |  |  |  |
| Female | 109<br>(52.9%) | 133 (42.6%) | 242 (46.7%) |

Additionally, a subset of participants also participated in a neuroimaging procedure, magnetic resonance imaging (MRI), where possible. While this study does not include this MRI data, the presence of this data may be indicative of a split in the participant group for testing the to-be developed bifactor model.

#### Data preparation

We selected different outcome variables per task based on three factors: (1) main outcome variables from test-batteries, (2) variables used in prior research (Brolsma et al., 2022; den Ouden et al., 2013; Heinzel, Northoff, Boeker, Boesiger, & Grimm, 2010; Janssen, Van Aken, De Mey, Witteman, & Egger, 2014; Mullane, Corkum, Klein, & McLaughlin, 2009; Reppermund, Ising, Lucae, & Zihl, 2009; Saylik & Szameitat, 2018; Stibbe, Huang, Paucke, Ulke, & Strauss, 2020), and (3) intercorrelations between outcome variables, including selecting only one of the variables from highly (R >.70) correlated sets. We inspected distributions by looking at scatterplots and histograms, and applied transformations in case of non-normality. We only excluded outliers (n=12) for one of the variables that showed extreme cases (IED: Total errors adjusted > 100). There were few data missing at random, therefore, we used Multiple Imputation by Chained Equations (MICE) predictive mean matching algorithm to correct for this (Wilson, 2021).

We standardized and split the cleaned dataset into two samples based on the availability of MRI data: discovery (N=206) and replication (N=312) (Table 1).

### Model Development and Replication

We used the Lavaan library in R to develop and estimate a bifactor structure underlying cognitive performance (Rosseel, 2012). We applied stepwise inclusion of the variables, evaluating the model fit iteratively (see supplementary materials S2).

We used the omega & Schmid Leiman transformation of the data, with a maximum likelihood estimation and oblique minimal rotation. For the general model fit, we considered the comparative fit index (CFI, good fit at > 0.95) and the root mean square error of approximation (RMSEA, acceptable fit between 0.01 and 0.08). These fit indices provide a broad sense of how well the model represents the data (Hu & Bentler, 1999). For the reliability of the general factor and subfactors of the model, we consider the hierarchical omega value, which is acceptable at >0.7 (Hu & Bentler, 1999; McNeish, 2018).

After stepwise inclusion of variables, count variables (e.g. Error variables) were excluded due to low contribution to the model fit. Eventually, both outcome variables from the probabilistic reversal learning task were excluded, resulting in a final 11-item model with CFI = 0.97, RMSEA = 0.09, Omega=0.57). This was the best fit, despite the omega index remaining below the commonly used threshold.

Replication of the model parameters in the replication set resulted in comparable model fit parameters, with CFI = 0.97, RMSEA = 0.05, and Omega = 0.55. Within both models, the eleven items load onto one converging C-factor, representing the shared variance over al included variables, and separately on 5 latent factors.

## Statistics

We used the bifactor model to characterize variation in CF in the sample. Given the absence of a priori directional hypotheses, associations were examined using group comparisons and partial correlations. All models were adjusted for age and sex, and statistical significance was corrected for multiple testing using the Holm-Bonferroni procedure.

First, we assessed whether differences in CF exist between broad categories of psychopathology - stress-related disorders (mood and anxiety disorders), neurodevelopmental disorders (ADHD / ASD and comorbid disorders (≥1 stress-related + ≥ 1 neurodevelopmental disorder) – using analysis of covariance (ANCOVA), followed by Tukey post-hoc pairwise comparisons.

Next, we explored the relation between CF in relation to specific diagnoses (mood-disorder, anxiety disorder, ADHD, ASC and addiction; coded 0 = absence, and 1 =presence of diagnosis) as well as total diagnostic burden (ranging from zero to five diagnoses), using partial Pearson correlation. Lastly, we also examined partial correlations between CF and dimensional symptom domains (Negative Valence, Cognition, Social systems and Inhibition/Arousal) as identified in the MIND-set cohort (Mulders et al., 2022).

In total 96 statistical tests were conducted: (6 group comparisons + 5 diagnoses + 1 comorbidity sum + 4 symptom dimensions) * 6 CF factors), with significance levels adjusted using *(ι/m)α* (with *m* = 96, *α* = 0.05).

## Results

This study identified and applied a bifactor model to assess the variability in CF within a naturalistic psychiatric patient sample.

### Bifactor Model

The Discovery and Replication model indicated that the eleven observed variables that were selected after step-wise inclusion, loaded onto similar factors (See Figure 1.A. and B). All eleven load onto one converging C-factor, representing the shared variance over all these variables, and separately on 5 latent factors: Reaction Time (RT), Incompatibility (IC), Working memory (WM), Inhibition (INH) and Flexibility (FLX).

**Figure 1.**
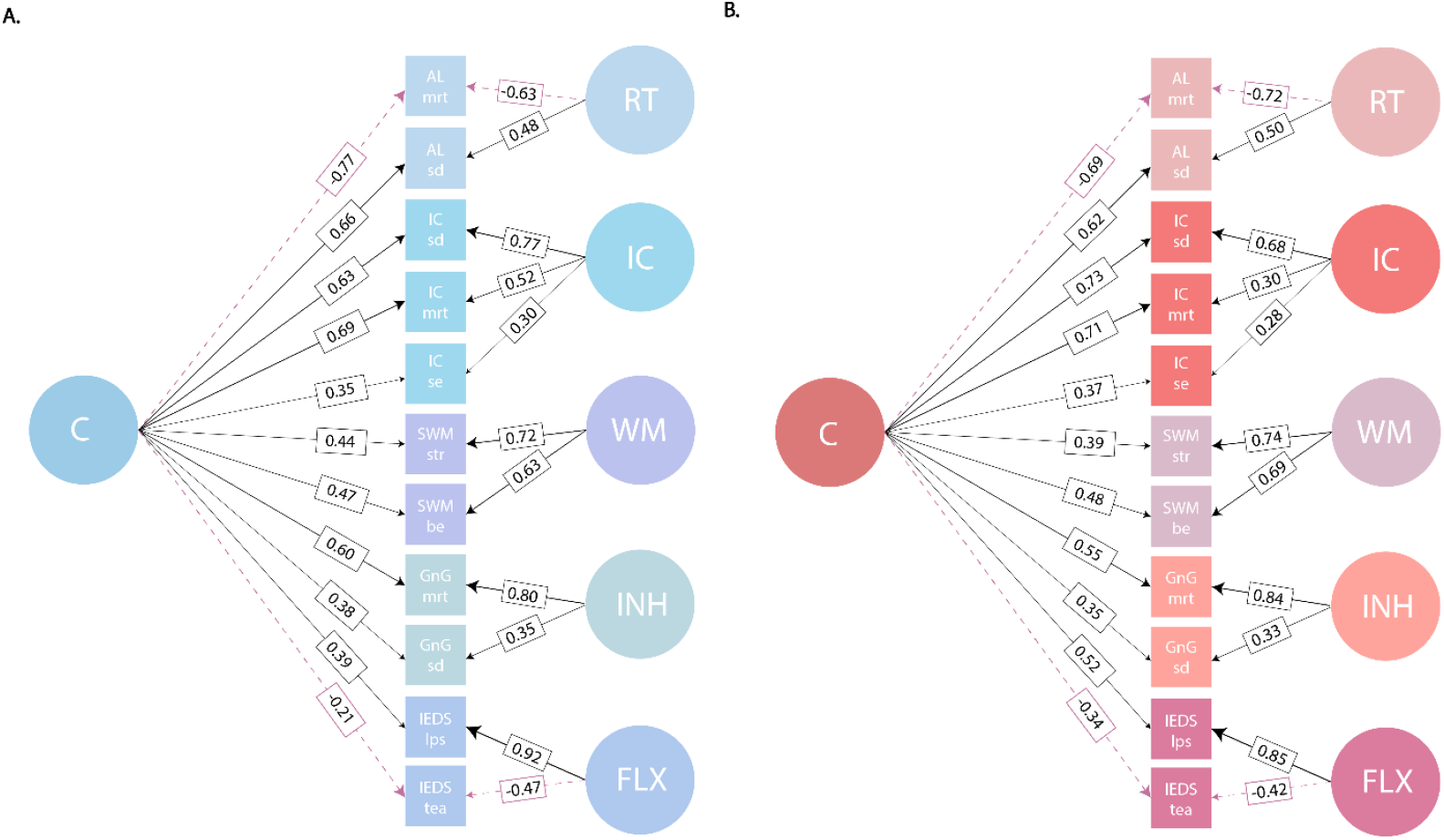
**A**. Bifactor model of cognitive functioning in patient sample without MRI data (N=206). The model is composed of 11 outcome variables from the task, that load onto one converging variable, the “C-factor” and separately on 5 latent factors we named according to the tasks: Reaction Time (RT), Incompatibility (IC), Working memory (WM), Inhibition (INH) and Flexibility (FLX). **B**. Bifactor model replicated using the same output variables and model parameters in the patient sample with MRI data (N=312). The model results in the same latent factors, with fit indices of RMSEA = 0.05 CFI = 0.97 Omega = 0.55.

#### Factor 1: Reaction Time - RT

Reaction Time included variables of the Alertness (AL) task, including the mean reaction time (mrt) and the standard deviation (sd). This task asks the participant to react as fast as possible to a stimulus appearing on the screen.

#### Factor 2: Incompatibility INC

Three variables from the incompatibility task load onto this factor. In this task participants needed to process divergent stimulus information, in parallel, requiring them to use a level of flexibility, impulse control and attentional selectivity (Zimmermann & Fimm, 2002). This concerned the appearance of an arrow that could point to, and appear at either the left or right. Participants had to press the key of the direction the arrow pointed in. The reaction time may be influenced if the direction of the arrow is incongruent with the location it appears in (e.g. arrow pointing to the left, appearing on the right). This effect is called the Simon Effect. The three variables that loaded onto the factor were the mean reaction time (mrt), standard deviation (sd) and the Simon Effect (se).

#### Factor 3: Working Memory – WM

The between errors (be) and the Strategy (str) variables from the Spatial Working Memory task, loaded onto this factor. This task requires participants to find tokens hidden in boxes on screen and measures visuospatial working memory. The *Between Errors* score is the number of mistakes made during the task, while *Strategy* score measures consistency in the approach of the task (Cambridge Cognition; Ltd, 2023)

#### Factor 4: Inhibition – INH

The variables mrt and sd from the Go-No Go task loaded onto the Inhibition factor. The Go-No Go task requires participants to respond quickly to a plus “+” sign appearing on the screen, but to not respond to a cross “x”.

#### Factor 5: Flexibility (FLX)

Two variables from the intra-extra dimensional set shift, loaded onto the final factor that we named Flexiblity. This task measures flexibility and set shifting abilities, by requiring participants to work out the rules that determines which stimulus is correct. It features multiple levels of difficulty. The variable included the Total errors adjusted (tea), which is the errors made in total adjusted by the total levels attained, and the latency per stage (lps), meaning the time taken per level of the task.

### Associations between CF and Psychopathology

#### Differences in CF between broader participant groups

The ANCOVA testing indicated only significant differences in the C-factor and the Incompatibility factor (INC) between broader participant groups (SD, ND, CD and control group) (figure 2.A). For both factors the SD-group scored significantly worse than the control group (C-factor p<0.0001, INC: p=0.0001). The four other factors did not show any significant between-group differences (all p>0.0005).

**Figure 2.**
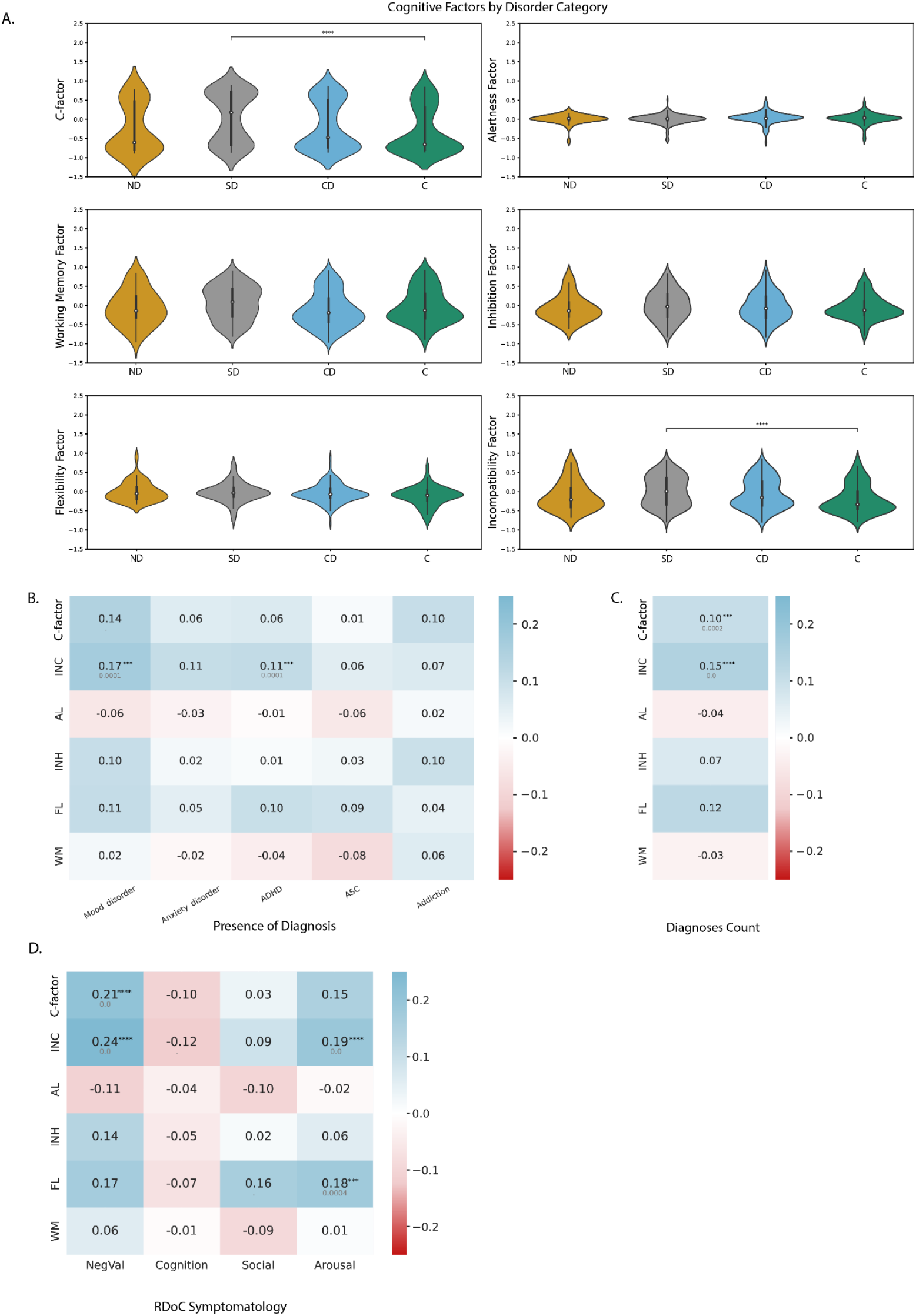
Overview of results describing the association between the bifactor model of CF and categories of psychopathology, significance is indicated with asterisks (with p <=0.005, ****: p<=0.0001, as due to multiple testing only p<0.005 is significant) With from top to bottom, left to right: A. the distribution per factor of the model, showing the difference per broader participant groups including Neurodevelopmental (ND), Stress-related (SD), Comorbid (CD), and non-disordered Controls (C). B. A partial correlation matrix with each of the CF factors correlated with the presence of a diagnosis (Mood or Anxiety disorder, ADHD, ASD and addiction), regardless of comorbidity C. A partial Pearson correlation matrix for each of the CF factors with the total count (0-5) of diagnoses. D. A Pearson correlation matrix for each of the CF factors with the dimensional symptom measures of severity (Negative Valence (NegVal), Cognition, Social and Arousal)

#### Association between CF and traditional diagnoses

As an initial exploratory analysis, we examined associations between the presence of individual psychiatric diagnoses and cognitive functioning (CF) using partial Pearson correlations adjusted for age and sex. After correction for multiple testing, a significant positive association was observed between the Incompatibility factor and the presence of a mood disorder (r = 0.17, p = 0.0001). No significant associations were found between any of the other CF factors, including the general factor, and the presence of the remaining diagnoses.

We next examined the association between cumulative diagnostic burden (total number of diagnoses, range 0–5) and CF using partial correlations adjusted for a age and sex and included in the same multiple-testing correction procedure (Figure 2C). A significant but weak positive correlation was observed between diagnostic burden and the Incompatibility factor (r = 0.19, p < 0.0001). Diagnostic burden was not significantly associated with any of the other CF factors.

Descriptive characteristics of the participant sample, including diagnostic combinations and corresponding scores on all bifactor CF dimensions, are presented in Table S3 of the Supplementary Materials.

#### Associations with transdiagnostic psychopathology

Lastly, we examined associations between CF and dimensional symptom domain scores, aligned with the RDoC framework, using partial correlations adjusted for age and sex and included in the multiple testing correction procedure (Figure 2.D). Significant associations were observed primarily with the Negative Valence domain. Specifically, higher Negative Valence scores were associated with greater impairment on the IC-factor (r = 0.24, p < 0.0001) and the C-factor (r = 0.20, p < 0.0001). Additionally, the IC factor and FL factor were significantly correlated with the Arousal dimension (IC: r=0.21, p < 0.0001, FL: r = 0.18, p = 0.0004). No significant associations were observed between any CF factors and the Social or Cognition domains.

## Discussion

In this study, we aimed to develop and evaluate a bifactor model of cognitive functioning based on data of a naturalistic cohort of patients and healthy controls. Then, we tested whether the resulting model relates to both traditional diagnoses and transdiagnostic dimensions of psychopathology. We found evidence for variable associations between psychopathology and the different factors of the model.

### Association of bifactor model with psychopathology

#### Associations based on diagnostic categories

We found the strongest associations between the general C-factor and the Incompatibility factor and diagnostic groups. A significant difference within these two factors existed between only the stress-related group and the non-disordered control group. Additionally, both the C-factor and the incompatibility factor correlated significantly with the total count of diagnoses, suggesting that poorer score in incompatibility related tasks and general CF functioning is linked to a higher diagnostic burden. However, contrary to expectations, higher impairment of cognition, indicated by a higher C-factor, was not present in the comorbid patient group. This is interesting as this group has the probability of a higher number of diagnoses. Notably, only the incompatibility factor was significantly associated with mood disorder diagnoses. Patient groups in which ADHD could be present (ND or CD group), did not differ significantly from other patient groups or the non-disordered group. More broadly, neither belonging to the ND or CD group nor having a diagnosis of ASC or anxiety appeared to be associated with significantly impaired CF. Taken together, this suggests that while a higher diagnostic burden could exacerbate CF dysfunction, this relationship is not strictly proportional, and may be more pronounced due to the presence of specific symptom profiles that are characteristic of a (severe) mood and/or ADHD diagnosis. This pattern points toward the potential relevance of transdiagnostic symptom dimensions, which we explore in the following section.

Scores on the Incompatibility task, or similar tasks, have earlier been found to be impacted in participants suffering from mood disorders and ADHD, and scores have also been negatively associated with sensation seeking and a lack of perseverance in non-disordered participants (Boxhoorn et al., 2019; Lamichhane, Moukaddam, Salas, Goodman, & Sabharwal, 2025). Earlier studies in this cohort indicated that similar levels of CF and ADHD symptomatology were present in patients with depression and ADHD, and comorbidity of both conditions (van Hal et al., 2023). These findings suggest that the incompatibility task or factor may reflect underlying cognitive traits common to both clinical and subclinical populations, aligning with evidence of overlapping symptomatology and cognitive profiles in individuals with depression, ADHD, or both. This overlap brings us to the transdiagnostic associations.

#### Transdiagnostic associations

The Incompatibility subfactor and the C-factor were significantly associated with the negative valence domain, while the Incompatibility and Flexibility factor were significantly associated with the arousal domain. In this context, arousal represents a transdiagnostic symptom domain encompassing impulsivity, antagonism, self-monitoring difficulties, and emotional distress, as measured by questionnaire items focused on personality, cognition, and ADHD-related symptoms. Given these characteristics, it is unsurprising that arousal was linked to Incompatibility and Flexibility. However, it was not associated with the Inhibition subfactor, which may be due to differences in task complexity. The inhibition task was a simple Go/No-Go paradigm, likely requiring fewer executive resources, whereas the Incompatibility and Flexibility tasks involved higher cognitive demands. This was also shown in a study by Boxhoorn and colleagues (2019) assessing basic attention and EF in families with and without ADHD. The increased cognitive load required for these tasks—and the resulting scores on their respective subfactors—may therefore be more sensitive to elevated psychopathology, particularly negative valence and arousal. It is worth considering that this relationship may follow an inverted u-shape rather than being linear, as is suggested by stress theories such as the Yerkes-Dodson framework (Beerendonk et al., 2024).

Earlier studies have shown that impairment of CF or EF was more associated with both general psychopathology as well as more specific symptoms dimensions, rather than specific diagnoses (Abramovitch et al., 2021; Bloemen et al., 2018). Furthermore, general psychopathology, or a P-factor, has been associated with impairments in CF: While in this study we did not assess general psychopathology in this way, the amount of diagnoses and transdiagnostic symptoms can indicate a broad level of severity and chronicity of psychopathology. Overall, the C-factor and the subfactor of incompatibility had the strongest associations, in both the diagnostic categories related to mood and ADHD, and the number of diagnoses. Additionally, these factors had significant associations with the transdiagnostic symptom domains related to negative valence and arousal, domains that are typically present in those with a mood-related or ADHD diagnosis. Accordingly, cognitive impairment may reflect the severity and transdiagnostic symptom burden of psychopathology rather than diagnostic classification alone, which may explain why such impairments are evident in some clinical groups or individuals but not consistently across all diagnoses. It is also important to note that natural variability in cognitive skills across the population may contribute to the observed associations, though this source of variation was not explicitly modeled.

Additionally, it is important to consider the discrepancy between self-reported and performance-based cognition. The multidimensional nature of Arousal as a transdiagnostic symptom domain, may not be fully captured by a single task, which could explain why its association with, for instance, inhibition was absent. Relatedly, the transdiagnostic symptom domain of Cognition showed only weak and negative correlations with all CF factors. This reinforces the idea that self-reported cognition and performance-based CF are only weakly, if at all, related. Instead, self-reports may be more influenced by personality traits or negative biases, as suggested by previous studies (Bodenburg, Wendiggensen, & Kasten, 2022; Buchanan, 2016; Snyder, Friedman, & Hankin, 2021). Taken together, these findings highlight the need for a nuanced interpretation of cognitive functioning in clinical research, emphasizing that performance-based and self-reported measures may tap into distinct yet complementary aspects of psychopathology.

### The bifactor model

Although the identified bifactor model had a good general fit (CFI, RMSEA), the hierarchical aspect of the model and reliability of the C-factor and subfactor index (omega) fell just below acceptable. In spite of that, we could replicate the precise constellation of subfactors with similar fit indices in a separate participant sample, indicating a consistency of the data. The subfactors replicate the original tasks on which we wanted to base the model, implying that our model mostly reflects task-specific variance, rather than more general cognitive constructs. The general C-factor is weak, while the within-task correlation is high, which may have caused the sub-factors to absorb the task-specific noise or method variance. The high level of heterogeneity in this participant sample may also influence the high variety in individual performance, allowing for a higher within-task variability. Further aspects that could have caused these task-specific subfactors are the selection of tasks (which may not share as much cognitive variance as expected), the selection of variables (as count-variables such as errors were excluded), and the fact that task-outcomes was based on one single performance. These tasks do not measure isolated cognitive processes in a strictly independent manner. Instead, they engage multiple interrelated domains of cognitive functioning. For instance, tasks that assess cognitive flexibility also require working memory and response inhibition, while those targeting reward processing involve error monitoring and adaptive decision-making. Additionally, it is important to recognize that the subfactors and general C-factor capture different aspects of the relationship between cognition and psychopathology. The C-factor encompasses the shared variance present in all the variables, while the subfactors resembles the unique variance of that task.

## Strengths & Limitations

This study has some strengths and limitations. Firstly, the main strength is that we have a naturalistic participant sample and could include a rich array of data including different symptom dimensions and performance based executive tasks. This allowed for a transdiagnostic take on cognitive function as a broader concept, rather than a more disorder-specific method.

Several methodological limitations may have shaped the structure of the bifactor model. The exclusion of variables from the Probabilistic Reversal Learning (PRL) task—likely due to its emotional feedback component and emphasis on “hot cognition”—narrowed the model’s scope. The weak C-factor and dominance of task-specific subfactors suggest limited shared cognitive variance across tasks, potentially compounded by reliance on single-session performance and the exclusion of error-based metrics. This design also prevented consideration of cognitive fluctuation, which may influence executive functioning across time (Judd, 2023). As a result, the model may have been less sensitive to broader cognitive constructs, instead reflecting task-specific variance more strongly than intended. In conclusion, impaired broader executive functioning, as captured by the C-factor, and tasks related to inhibition with a higher cognitive load are mostly associated with mood-disorders as well as symptoms of negative valence and arousal. Our findings highlight the importance of considering both general and task-specific cognitive performance, as well as transdiagnostic symptoms when assessing executive dysfunction in clinical populations.

## Data Availability

All data produced in the present study are available through formal request to the authors

https://www.ru.nl/en/donders-institute/research/di-funded-research-projects/research-projects/the-mind-set-project

