## Supplementary Materials for "The Transdiagnostic Association between Cognitive Functioning and Psychopathology *Exploratory Modeling of Cognitive Structure in a Naturalistic Patient Sample*"

*Table S1. Selected variables per test battery and task*

| Test Battery | Task | Measures | Observed variable | Selected variables - abbreviation |
|---|---|---|---|---|
| **TAP** | Alertness | Basal responsiveness, general processing speed, reaction stability, differentiation tonic and phasic alertness. | • Mean reaction Time (No Alarm)<br>• Standard Deviation (No Alarm)<br>• Anticipation Errors (Alarm)<br>• Phasic Alertness | • Mean reaction Time (No Alarm) AL-mrt<br>• Standard Deviation (No Alarm) – AL-sd<br>• Phasic Alertness - AL-pha |
| | Go- no Go | Inhibition ability, impulse control | • Errors<br>• Omissions<br>• *Mean Reaction Time*<br>• *Standard Deviation* | • Mean reaction Time (No Alarm) GnG - mrt<br>• Standard Deviation – GnG-sd<br>• |
| | Incompatibility | Cognitive flexibility, handling of cognitive interference | • Errors<br>• *Omissions*<br>• *Mean Reaction Time*<br>• *Standard Deviation*<br>• Simon Effect (general) | • Mean reaction Time – INC- mrt<br>• Standard Deviation – INC – sd<br>• Simon Effect – INC -se |
| **CANTAB** | Intra-Extra Dimensional Set Shift (IEDS) | Cognitive flexibility/Set-shifting, | • Total Errors Adjusted<br>• Latency per Stage | • IEDS – tea<br>• IEDS - lps |
| | Spatial Working Memory (SWM) | Working memory, short term memory | • Between Errors<br>• Choice Duration<br>• Strategy | • SWM- be<br>• SWM – cd<br>• SWM - str |
| **PRL** | Probabilistic Reversal Learning | Probabilistic reasoning, reward learning | • Perseverative Errors<br>• Probabilistic Switch Rate (PSR) | • PRL – pe<br>• PRL - psr |

*Table S2. Fit indices per step of included variables.*

| STEP | INCLUDED VARIABLES | RMSEA | CFI | OMEGA |
|---|---|---|---|---|
| 1 | GnG-mrt, GnG-err, IC-se, IC-mr, SWM-str, AL-mrt, AL-fa, IEDS-tea | 0.07 | 0.96 | 0.06 |
| 2 | GnG-mrt, GnG-err, IC-se, IC-mr, IC-err, SWM-str, AL-mrt, AL-fa, IEDS-tea, SWM-be, PRL-pe | 0.09 | 0.91 | 0.22 |
| 3 | GnG-mrt, GnG-err, IC-se, IC-mr, IC-err, SWM-str, AL-mrt, AL-fa, IEDS-tea, SWM-be, PRL-pe, PRL-psr | 0.03 | 0.95 | 0.13 |
| 4 | GnG-mrt, GnG-err, GnG-sd, IC-se, IC-mr, IC-err, SWM-str, AL-mrt, AL-fa, IEDS-tea, SWM-be, PRL-pe, PRL-psr | 0.02 | 0.96 | 0.17 |
| 5 | GnG-mrt, GnG-err, GnG-sd, IC-se, IC-mr, IC-sd, SWM-str, AL-mrt, AL-fa, AL-sd, IEDS-tea, SWM-be, PRL-pe, PRL-psr | 0.12 | 0.97 | 0.57 |
| 6 | GnG-mrt, GnG-sd, IC-se, IC-mrt, IC-sd SWM-str, SWM-be, AL-mrt, AL-sd, IEDS-tea, IEDS-lps, PRL-psr | 0.11 | 0.92 | 0.53 |
| 7 | GnG-mrt, GnG-sd, IC-se, IC-mrt, IC-sd SWM-str, SWM-be, AL-mrt, AL-sd, IEDS-tea, IEDS-lps | 0.09 | 0.97 | 0.57 |
| 8 | GnG-mrt, GnG-sd, IC-se, IC-mrt, IC-sd SWM-str, SWM-be, AL-mrt, AL-sd, IEDS-lps | 0.11 | 0.96 | 0.57 |

*Table S3. Separate factor scores per diagnosis combination.*

| Diagnosis Combination | N | C-factor | IC_fac | AL_fac | INH_fac | FL_fac | WM_fac |
|---|---|---|---|---|---|---|---|
| Controls | 112 | -0.71 | -0.45 | 0.04 | -0.19 | -0.23 | -0.04 |
| Addiction | 3 | -0.89 | -0.16 | 0.12 | -0.05 | -0.75 | -0.65 |
| ASC | 14 | -0.8 | -0.5 | 0.01 | -0.22 | -0.1 | -0.04 |
| ASC_Addiction | 5 | 0.66 | 0.66 | -0.01 | 0.46 | -0.48 | -0.39 |
| ADHD | 18 | -0.66 | -0.25 | 0.0 | -0.26 | 0.06 | -0.19 |
| ADHD_Addiction | 8 | 0.53 | -0.07 | 0.23 | 0.31 | -0.01 | 0.05 |
| ADHD_ASC | 7 | -1.26 | -0.52 | 0.03 | -0.31 | -0.01 | -0.77 |
| ADHD_ASC_Addiction | 2 | -0.5 | 0.03 | 0.09 | 0.19 | -0.42 | -1.17 |
| Anxiety_disorder | 8 | -0.09 | 0.05 | -0.08 | -0.42 | 0.06 | 0.49 |
| Anxiety_disorder_Addiction | 1 | 0.57 | -0.1 | 0.04 | 0.57 | 0.05 | 0.38 |
| Anxiety_disorder_ASC | 4 | 0.1 | 0.39 | -0.76 | 0.53 | 0.3 | -0.01 |
| Anxiety_disorder_ASC_Addiction | 0 | | | | | | |
| Anxiety_disorder_ADHD | 3 | -0.4 | 0.15 | 0.15 | -0.13 | 0.05 | -0.98 |
| Anxiety_disorder_ADHD _Addiction | 2 | 1.85 | 0.44 | 0.8 | -0.13 | -0.09 | 0.38 |
| Anxiety_disorder_ADHD _ASC | 1 | -0.48 | 0.16 | 0.09 | -0.74 | -0.26 | 0.19 |

| | | | | | | | |
|---|---|---|---|---|---|---|---|
| Anxiety_disorder_ADHD_ASC_Addiction | 0 | | | | | | |
| Mood_disorder | 73 | 0.53 | 0.27 | -0.01 | 0.15 | 0.02 | 0.18 |
| Mood_disorder_Addiction | 31 | 0.18 | -0.01 | 0.0 | 0.13 | 0.05 | 0.05 |
| Mood_disorder_ASC | 22 | -0.79 | -0.52 | -0.04 | 0.08 | -0.23 | -0.25 |
| Mood_disorder_ASC_Addiction | 4 | -1.33 | -0.52 | 0.2 | -0.35 | -0.18 | -0.96 |
| Mood_disorder_ADHD | 35 | 0.07 | 0.19 | 0.07 | -0.09 | -0.12 | -0.11 |
| Mood_disorder_ADHD_Addiction | 16 | 1.06 | 0.6 | -0.2 | 0.35 | 0.46 | 0.17 |
| Mood_disorder_ADHD_ASC | 16 | -0.68 | -0.1 | 0.01 | -0.35 | -0.07 | -0.4 |
| Mood_disorder_ADHD_ASC_Addiction | 8 | 0.61 | 0.12 | -0.1 | 0.28 | -0.08 | 0.64 |
| Mood_disorder_Anxiety_disorder | 38 | 1.3 | 0.83 | -0.05 | 0.34 | 0.07 | 0.28 |
| Mood_disorder_Anxiety_disorder_Addiction | 7 | 0.83 | -0.0 | 0.12 | 0.47 | -0.07 | 0.47 |
| Mood_disorder_Anxiety_disorder_ASC | 24 | -0.61 | -0.17 | 0.01 | 0.02 | 0.13 | -0.77 |
| Mood_disorder_Anxiety_disorder_ASC_Addiction | 5 | 0.39 | 0.32 | 0.0 | -0.18 | -0.01 | 0.3 |
| Mood_disorder_Anxiety_disorder_ADHD | 12 | -0.56 | -0.19 | -0.09 | 0.01 | -0.32 | -0.11 |
| Mood_disorder_Anxiety_disorder_ADHD_Addiction | 6 | -1.27 | -0.24 | 0.15 | -0.64 | -0.48 | -0.6 |
| Mood_disorder_Anxiety_disorder_ADHD_ASC | 7 | -1.5 | -0.3 | 0.16 | -0.97 | -0.12 | -0.74 |
| Mood_disorder_Anxiety_disorder_ADHD_ASC_Addiction | 1 | -2.16 | -0.99 | 0.17 | -0.7 | -0.09 | -0.6 |
